# A Cross Sectional Study To Assess The Attitude Regarding Online Lecture Among Nursing College Students After Impact of Covid-19 At Selected College of Nursing, Nadiad

**DOI:** 10.1101/2021.07.01.21259132

**Authors:** Kailash Nagar

## Abstract

**Background:** In December 2019, a pathogenic human corona virus SARS- CoV-2, corona virus disease 2019 (COVID-19), was recognized and has caused serious illness and numerous deaths. The epidemics of COVID-19 have been recorded over 200 countries, territories, and areas with 2 878 196 confirmed cased and 198 668 death cases.

**Aims:** The current study is to assess the attitude of nursing college students regarding online classes.

**Objective:** **1**. To determine the perceptions of students towards e-learning during the lock down. 2. To assess nursing students attitude towards the online lectures

**Methodology:** *Setting and Design:* Descriptive cross sectional survey research design was used and not probability sampling methods was used to drawn samples through online Google form. The sample collected from nursing college students and study setting was Dinsha Patel College of nursing, Nadiad, Gujarat. There were total 20 attitude question with five point rating scale were used. Total sample were 136 nursing college students. Data has been collected online through Google form.

*Statistical Analysis used:* Descriptive statistics applied where, data was analyzed by using SPSS software, and Frequency, percentage, tables etc were used to represent the statistical data.

**Results:** Out of 136, 64(47.05%) were belong age 17-18 years, 115(84.56%) were female, 68(50%) were GNM students, 51(37.50%) B.Sc. nursing Students, 83(61.02%) were third year, 75(55.24%) were live urban, 61(44.86%) were rural area, 56(41.17%) were travel 21-30km, 40(29.41%) more than 30km, 65(47.79%) have above 15000 monthly family income 132(97.05%) were used mobile for online lecture 79(58.08%) were have average network.

Attitude regarding Online Classes where 7 (5.14%) had Inadequate Attitude 86(63.25%) had Moderate Attitude, 43(31.61%) Had adequate Attitude.

**Conclusions:** Regarding the online lecture 63.25% nursing college students has moderate attitude and 31.61% had adequate attitude it means now days students are line online classes than physical mode due to various reasons such as, time consuming, reduce traveling, risk of accidents and corona, more time can spend with family etc

## Introduction: - “The beautiful thing about learning is that nobody can take it away from you”

On 11 March 2020, WHO changed the status of the COVID-19 emergency from public health international emergency (30th January 2020) to a pandemic. Due the COVID-19 pandemic situation college students are not able to join college physically and as per WHO guideline all has to remain at home and maintain social distance, thus students has to take classes from their home. But it’s very difficult to all the students to attend classes from home they may faced certain problems such as bad network, financial problems, not availability of internet, slow speed of internet etc.

Online learning is a need of the day and in India, The government and other educational organizations had design policies for online learning to implement in education field at university level, but due to less knowledge about ground realties, the policies fails to achieve the desired outcomes. A number of researches were made on various dimension of online learning, i.e., barriers and impact of online learning in the students’ academic achievements, but least study was made on the attitude of students towards online learning. Moreover, the studies were made at secondary and intermediate level but no formal efforts were made to conduct research at undergraduate level. As the undergraduate level is a crucial stage in academics and the students are more motivated and energetic at this level to perform tangible research work, therefore, to know their attitude regarding online learning is of great importance.

## OBJECTIVES

The purpose of this study was

1. To determine the perceptions of students towards e-learning during the lock down.
2. To assess nursing students attitude towards the online lectures

## RESEARCH METHODOLOGY

### Research Approach

Non Experimental, Descriptive survey approach

### Research Design

Cross sectional study.

### Research Variables

1. **Dependant variables:** Attitude of Nursing Student on Online Lectures or Classes.
2. **Demographic variables:** demographic variables of Nursing Student’s such as Age, Gender, Course, Year, Distance of college from home, income of the family, Locality and type of family, Network Availability etc.

### Sampling method

The survey was prepared online and hyperlink of the survey was distributed to students using mobile group messaging application. It was made sure in a class that most of the students are having smart mobile devices and sufficient Internet connectivity to fill up the form online. Students who were not using Internet were encouraged to take help from their friends having Internet enabled device. Prior to the distribution, students were made clear about the objectives of this study and were also demonstrated an online learning system they expected to enroll in during the semester. It is to be noted that student participation was voluntarily and they could opted not to fill up the survey.

### Instrument for Data Collection

A closed questionnaire with 5-points Likert scale. Questionnaire was accompanied with many items, which were subjected for the collection of data from the respondents.

### Study population

All Nursing College Students. (GNM, B.Sc., P.B.B.Sc., M.Sc.)

### Study Sample

Nadiad, Nursing College students

### Study Setting

Dinsha Patel College of Nursing, Nadiad.

### Sample Size

136 Nursing College Student.

## SAMPLE CRITERIA

### Inclusion criteria

1. Nursing College Students of both sexes of age between 19-36 years,
2. Who have the facilities of internet in phone, Laptop and computer.
3. Those who willing to participate in study

### Exclusion criteria

1. Those who are not willing to participate in the research study.
2. The Students who haven’t the Facilities of smart phone or internet.

## TOOL FOR DATA COLLECTION

### Section-I

It Consist of demographic variables of Nursing Student’s such as Age, Sex, Course, Year, income of the family, Locality and type of family, etc.

### Section-II

It consists of structured self administered questionnaire (5-points Likert scale) to assess the nursing student’s attitude towards the online lectures or classes.

Table no.2 Revealed that the distribution of sample according to attitude regarding Online Classes were 7 (5.14%) had Inadequate attitude 86(63.25%) had Moderate attitude, 43(31.61%) Had adequate attitude.

**Table No.1.**
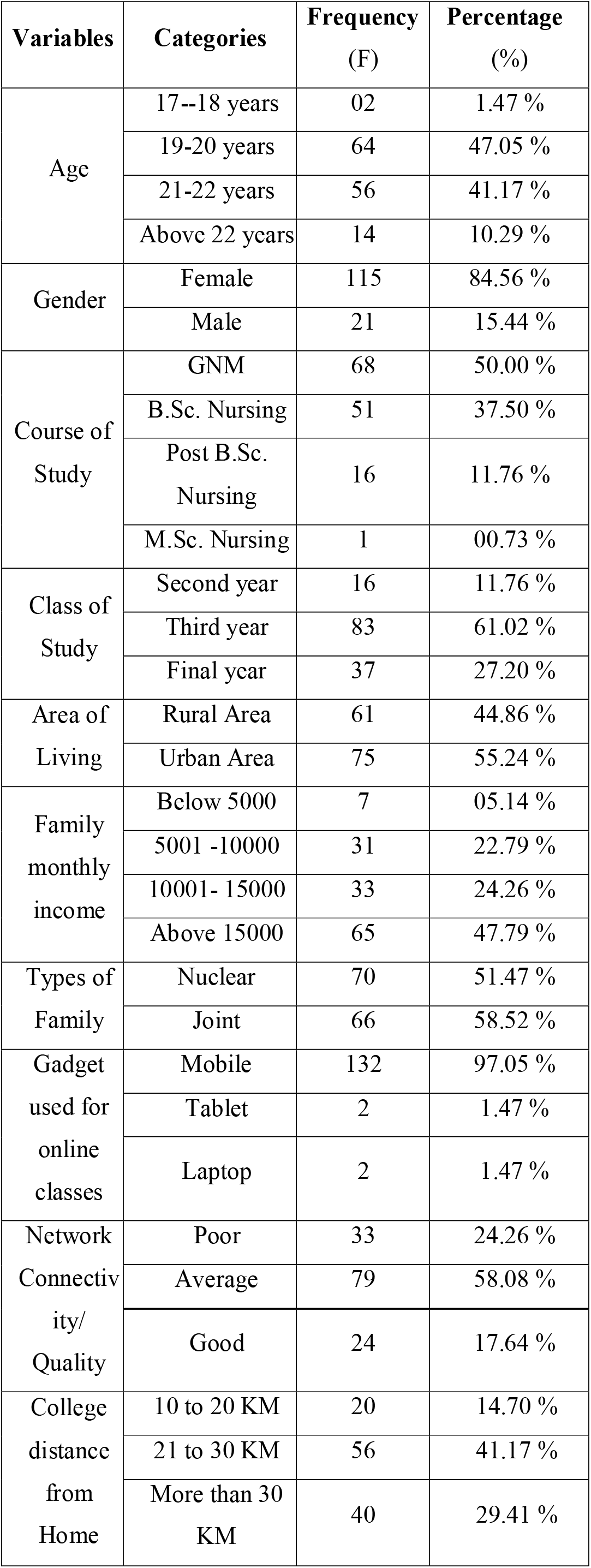
Demographic variable of nursing College students.

**Demographic variable of nursing College students.**
| Variables | Categories | Frequency (F) | Percentage (%) |
| --- | --- | --- | --- |
| Age | 17--18 years | 02 | 1.47 % |
|  | 19-20 years | 64 | 47.05 % |
|  | 21-22 years | 56 | 41.17 % |
|  | Above 22 years | 14 | 10.29 % |
| Gender | Female | 115 | 84.56 % |
|  | Male | 21 | 15.44 % |
| Course of Study | GNM | 68 | 50.00 % |
|  | B.Sc. Nursing | 51 | 37.50 % |
|  | Post B.Sc. Nursing | 16 | 11.76 % |
|  | M.Sc. Nursing | 1 | 00.73 % |
| Class of Study | Second year | 16 | 11.76 % |
|  | Third year | 83 | 61.02 % |
|  | Final year | 37 | 27.20 % |
| Area of Living | Rural Area | 61 | 44.86 % |
|  | Urban Area | 75 | 55.24 % |
| Family monthly income | Below 5000 | 7 | 05.14 % |
|  | 5001 -10000 | 31 | 22.79 % |
|  | 10001- 15000 | 33 | 24.26 % |
|  | Above 15000 | 65 | 47.79 % |
| Types of Family | Nuclear | 70 | 51.47 % |
|  | Joint | 66 | 58.52 % |
| Gadget used for online classes | Mobile | 132 | 97.05 % |
|  | Tablet | 2 | 1.47 % |
|  | Laptop | 2 | 1.47 % |
| Network Connectivity/ Quality | Poor | 33 | 24.26 % |
|  | Average | 79 | 58.08 % |
|  | Good | 24 | 17.64 % |
| College distance from Home | 10 to 20 KM | 20 | 14.70 % |
|  | 21 to 30 KM | 56 | 41.17 % |
|  | More than 30 KM | 40 | 29.41 % |

**Table No.2.** Distribution according to level of attitude.

| Attitude | Frequency (F) | Percentage (%) |
| --- | --- | --- |
| Unfavorable Attitude | 07 | 5.14 % |
| Moderate Unfavorable Attitude | 86 | 63.25 % |
| Favorable Attitude | 43 | 31.61 % |
| **TOTAL** | 136 | 100 % |

**Graph 01:**
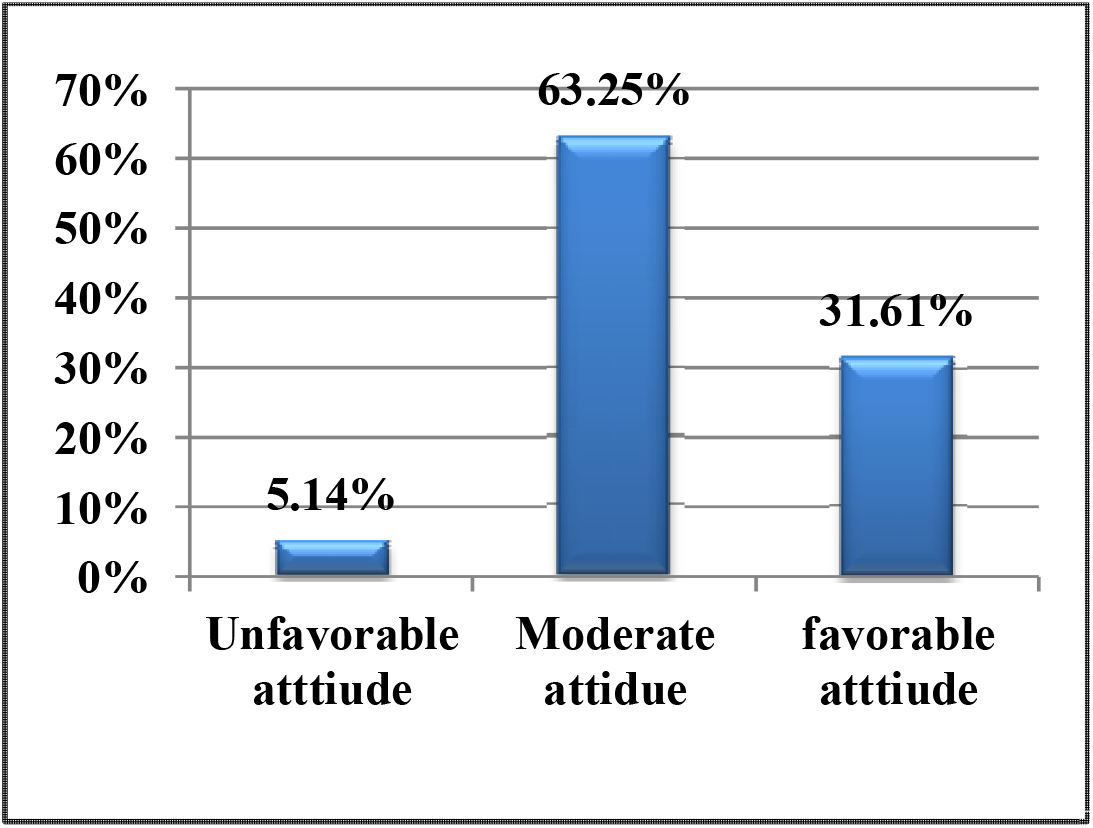
Distribution of Attitude toward online classes.

**Table No.3.**
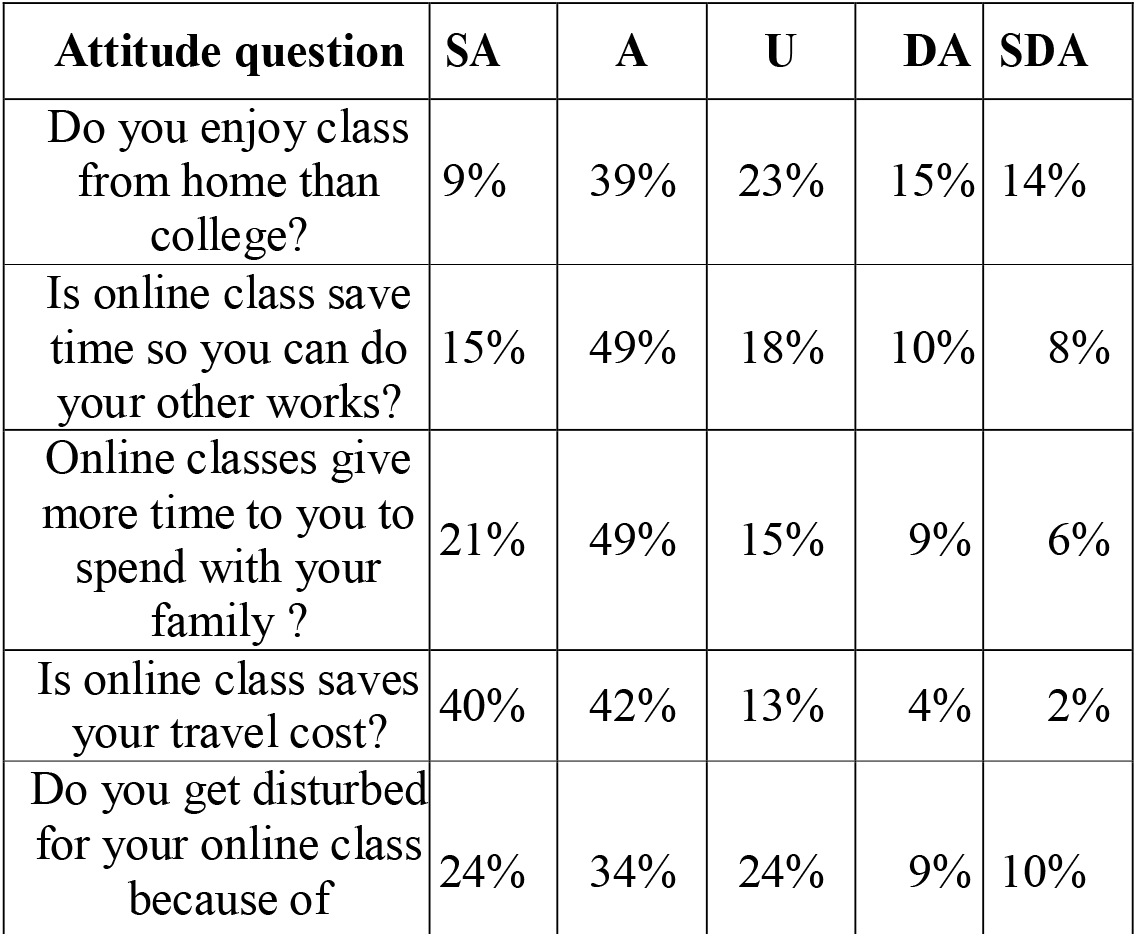
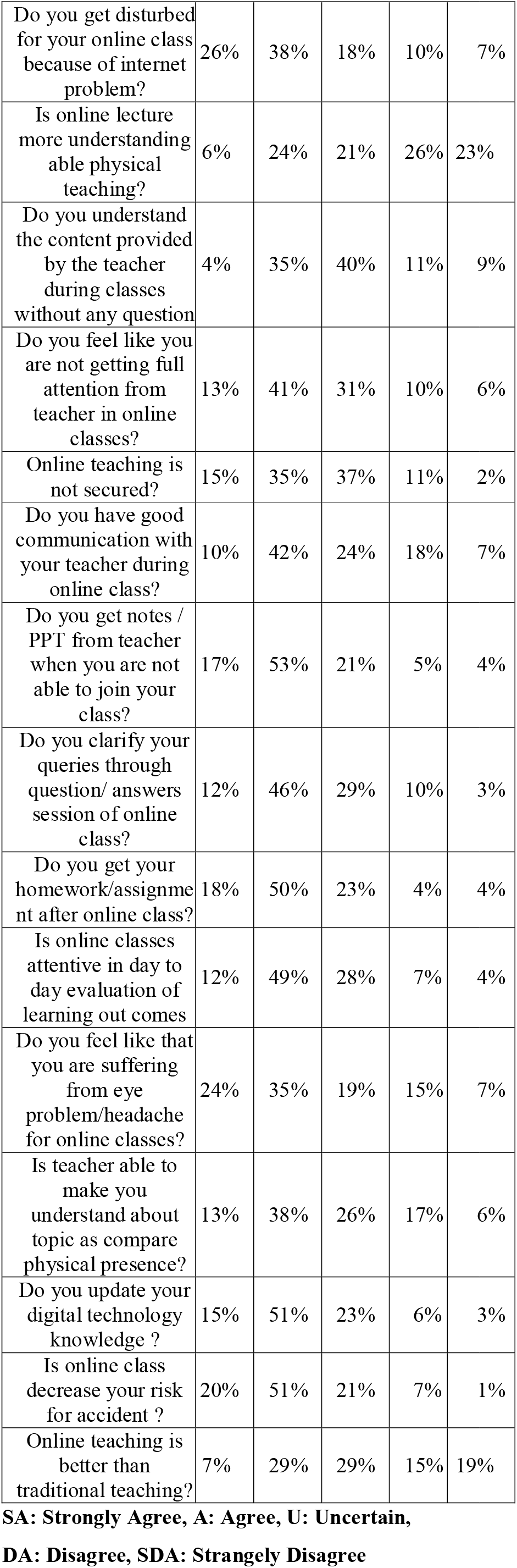
Distribution according to percentage of attitude questionnaire.

## Conclusion

On the basis of findings of this study the following conclusion were drawn:

The purpose of the present study is to assess the attitude regarding online lecture after the impact of COVID-19 at selected nursing college Nadiad. The study consisted of 136 samples that were selected on the basis of simple randomization techniques. Based on the objective, the data analysis was done by calculating the mean, percentage, standard deviation. Result revealed that an attitude of nursing students towards Regarding the choice of gadgets to attends online class of students out of 136 samples, 132(97.05%) was mobile, 2(1.47%) was laptop2(1.47%). Regarding the network quality of students out of 136 samples, 33(24.26%) was poor, 79(58.08%) was average, 24 (17.64%) good. Revealed that the distribution of sample according to Attitude regarding Online Classes were 7 (5.14%) had Inadequate Attitude 86(63.25%) had Moderate Attitude, 43(31.61%) Had adequate attitude.

## Data Availability

yes all data were included properly

## Conflict of Interest

Nil

## Source of Funding

College Management

## Ethical Clearance

The study was approved by the research committee, Institutional ethical Committee (IEC) – DPCN/1^st^ IEC/2018-19/09 and a formal written permission was gathered from the authority of Principal of Institute.

## Statement of Informed consent

Informed consent was acquired from the participants

---

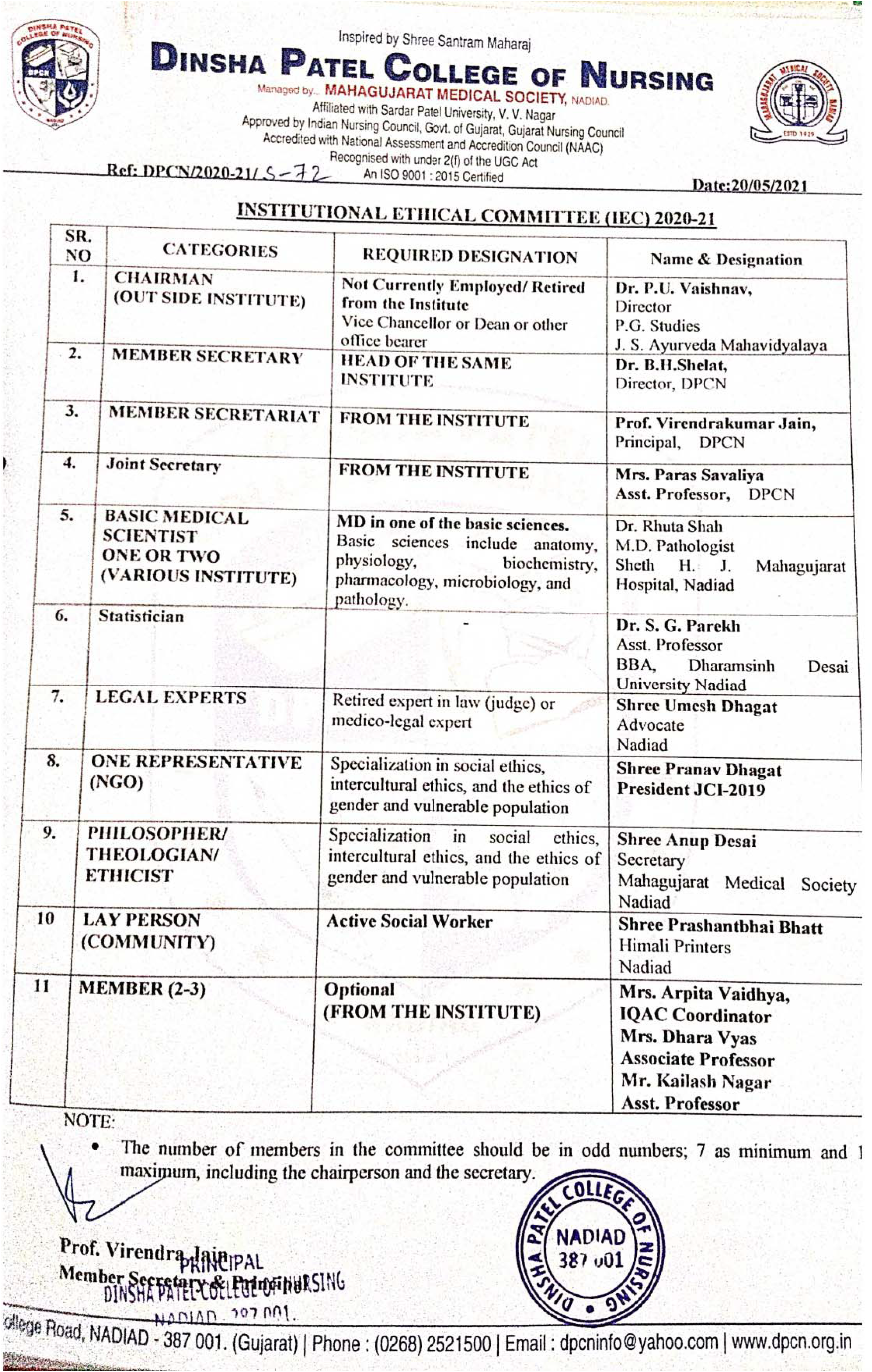

## Notes

### Competing Interest Statement

The authors have declared no competing interest.

### Clinical Trial

Clinical trial was not required because its an descriptive study and not any harm to subjects

### Funding Statement

self funded

### Author Declarations

The study was approved by the Institutional ethical Committee of Dinsha Patel College of Nursing, Nadiad. the Refence number of the ethical approval was DPCN 1st IEC 2018 and 19,09, there are more than 11 members of the ethical committee who were approved the study. And a formal written permission was gathered from the authority of Principal of Institute before conducting the study, also researcher has conducted consent form from the participants.

## REFERENCES

[1] Wentling, T. L., Waight, C., Gallaher, J., La Fleur, J., Wang, C., & Kanfer, A., “E-learning: A review of literature”. Urbana-Champaign: University of Illinois, 2000

[2] Rosenberg, Marc Jeffrey. “E-learning: Strategies for delivering knowledge in the digital age”. Vol. 9. New York: McGraw-Hill, 2001.

[3] Chorng-Shyong, Jung-Yu Lai, and Yi-Shun Wang. “Factors affecting engineers’ acceptance of asynchronous elearning systems in high-tech companies.” Information & management 41.6 (2004): 795–804.

[4] Exploring the Pros and Cons of Online, Hybrid and Face-to-face Class Formats. A Provost Report Series. University of Washington. 2013.

[5] Sanders, Diana W., and Alison I. Morrison-Shetlar. “Student attitudes toward web-enhanced instruction in an introductory biology course.” Journal of Research on computing in Education 33.3 (2001): 251–262

[6] Kailash Nagar, Knowledge, Attitude and Practice on Personal Hygiene among School Children in Rural Primary School of Kheda District, Gujarat. Indian Journal of Forensic Medicine & Toxicology [Internet]. 2021May17,15(3):290–5. Available from:http://medicopublication.com/index.php/ijfmt/article/view/15321

[7] Ali, Naila, et al. “Attitude of nursing students towards e-learning.” Advances in Health Professions Education 2.1 (2016): 24–29. International Journal of Advance Engineering and Research Development (IJAERD) Volume 4, Issue 11, November-2017, e-ISSN: 2348 -4470, print-ISSN: 2348-6406 @IJAERD-2017, All rights Reserved 213

[8] Dhamija, Neelam. “Attitude of Undergraduate Students Towards the use of e-Learning.” MIER Journal of Educational Studies, Trends and Practices 4.1 (2016): 123–125.

[9] Varshney, Anant Kumar. “Attitude of Rural and Urban Undergraduate Students of Aligarh Muslim University towards Computer.” EDUCARE 8.1 (2016): 97–104.

[10] Suri, Gunamala, and Sneha Sharma. “The impact of gender on attitude towards computer technology and e-learning: An exploratory study of Punjab University, India.” International Journal of Engineering Research 2.2 (2013): 132–136.

[11] Kailash Nagar. A Study to Assess the Effectiveness of Planned Teaching Programme on Prevention of Selected Life Style Diseases in Terms of Knowledge and Attitude among Male Adults at Selected PHC of Kheda District. Indian Journal of Forensic Medicine & Toxicology [Internet].2021May17, 15(3):2812–5. Available from: http://medicopublication.com/index.php/ijfmt/article/view/15732.

